# Therapeutic potential of a modified Mediterranean ketogenic diet in reversing the peripheral lipid signature of Alzheimer’s disease

**DOI:** 10.1101/2023.06.13.23291049

**Authors:** Bryan J Neth, Kevin Huynh, Corey Giles, Tingting Wang, Natalie A Mellett, Thy Duong, Colette Blach, Leyla Schimmel, Thomas C Register, Kaj Blennow, Henrik Zetterberg, Richa Batra, Annalise Schweickart, Amanda Hazel Dilmore, Cameron Martino, Matthias Arnold, Jan Krumsiek, Xianlin Han, Pieter C Dorrestein, Rob Knight, Peter J Meikle, Suzanne Craft, Rima Kaddurah-Daouk

## Abstract

Alzheimer’s disease (AD) is a major neurodegenerative disorder with significant environmental factors, including diet and lifestyle, influencing its onset and progression. Although previous studies have suggested that certain diets may reduce the incidence of AD, the underlying mechanisms remain unclear. In this randomized crossover study of 20 elderly adults, we investigated the effects of a modified Mediterranean ketogenic diet (MMKD) on the plasma lipidome, analyzing 784 lipid species across 47 classes. We identified substantial changes in response to intervention, notably a global elevation across all plasmanyl and plasmenyl ether lipid species among others, with many changes linked with changes to clinical and biochemical markers of AD. Leveraging our prior clinical studies on AD (n = 1,912), we found that MMKD was inversely associated with the lipidomic signature of prevalent and incident AD. Given its low risk and cost, MMKD could be a promising approach for prevention or early symptomatic treatment of AD.

## INTRODUCTION

Alzheimer’s disease (AD) is the most common neurodegenerative disorder and has an increasing incidence.^1^ Despite extensive research, efforts to identify clinically meaningful disease-modifying therapies have been difficult,^2–4^ making AD one of our most significant health challenges. The pathophysiologic hallmarks of AD occur long before the onset of clinical symptoms.^5^ As such, there is great interest in understanding the brain and systemic changes that occur in patients at risk for AD in hopes of preventing the development of progressive neurologic decline.^6^

The traditional ketogenic diet (KD) was developed at the Mayo Clinic in 1921 for the treatment of epilepsy, ^7, 8^ and continues be a part of clinical care for patients who have medically intractable epilepsy.^9^ KD is a high fat and low carbohydrate diet. This transition of the body’s primary metabolic fuel from glucose to fats and ketones leads to ketosis, or the production of ketone bodies.^10^ The KD has proven to be highly effective in the control of seizures, which has led to its continued clinical use.^9, 11^ A modified KD with the ability to sequentially increase carbohydrates has shown similar efficacy in seizure control,^12^ which improves long-term adherence.^13^

There are clear metabolomic changes in AD, highlighted by altered phospholipid and sphingolipid metabolism which begin in preclinical^14^ and symptomatic AD.^15^ Changes have also been noted in bile acids, triglycerides and lipoproteins, in cholesterol metabolism and clearance through bile acids, and in ether lipids including plasmalogens, branched chain amino acids and acylcarnitines^14–17^. Our Alzheimer’s Disease Metabolomics Consortium under the Accelerating Medicine Partnership for Alzheimer’s Disease has generated large data sets of metabolomics and lipidomics from large AD and community studies that defined many of these changes, and in which peripheral changes were connected with imaging changes and amyloid, tau and neurodegeneration markers of disease.^18–29^

Lipids are fundamental components of cellular structure and function, particularly in the brain, which is one of the most lipid-rich organs.^30^ Lipids are a major constituent of membranes and synapses,^31^ which are impaired and ultimately lost throughout the course of AD.^31, 32^ Thus, studying lipids in the context of preclinical and early symptomatic AD (mild cognitive impairment or MCI) provides an important window into crucial lipid changes and how they may be modified by intervention prior to advanced neuropathologic changes.

Lipidomics is a powerful tool used to study the diversity of multiple lipid classes and species.^33^ We have previously used a targeted lipidomics platform to map the key plasma lipid signatures in MCI and AD dementia across two large clinical cohorts, and found concordant changes in ether lipids, sphingolipids, triglycerides, and phosphatidylethanolamine.^34^ The lipid aberrations in AD serve as an important therapeutic target, particularly as many of the affected lipid classes have structural and functional roles in the plasma membrane involving signaling and synaptic function.^31, 32, 35–38^ The importance of lipids in AD is highlighted by *APOE*, a lipoprotein gene. *APOE* remains the largest genetic risk factor for sporadic AD, with the □4 isoform dramatically increasing the risk of AD.^39, 40^ Conversely, the APOE □*2* genotype reduces the risk of AD, and we have previously identified that around one-third of the protective effects of *APOE* □*2* are potentially mediated through peripheral ether lipids^41^.

Here we use a robust targeted lipidomics platform in which 784 species across 47 classes are measured to map the effect of a KD on the plasma lipidome in patients at risk for AD with and without cognitive impairment (Supplementary Table 9). Our goals are to address the following questions: (1) How does the KD change the lipidome; (2) How do the lipidome changes from a KD compare to those from a low-fat diet; (3) Are lipidome changes after a KD different between clinical subgroups; and (4) Can the KD act as a therapeutic by reversing the AD lipid signature?

## RESULTS

### The modified Mediterranean ketogenic diet significantly altered the plasma lipidome at a class level

Lipidomic measures were taken on all plasma samples collected in the study. We examined the impact of the MMKD and AHAD on the plasma lipidome using linear mixed models after each dietary intervention. Significant impacts on the plasma lipidome were observed in the MMKD (Figure 1A) with 18 of the 47 categorized lipid classes presenting an association with an FDR corrected p-value < 0.05. In contrast, the AHAD had no associations after FDR correction, but 12 lipid classes presented with nominal associations with uncorrected p < 0.05 (Supplementary Table 1). There were minimal impacts to the lipidome after washout periods relative to pre-study baseline (Supplementary Figure 2).

**Figure 1:**
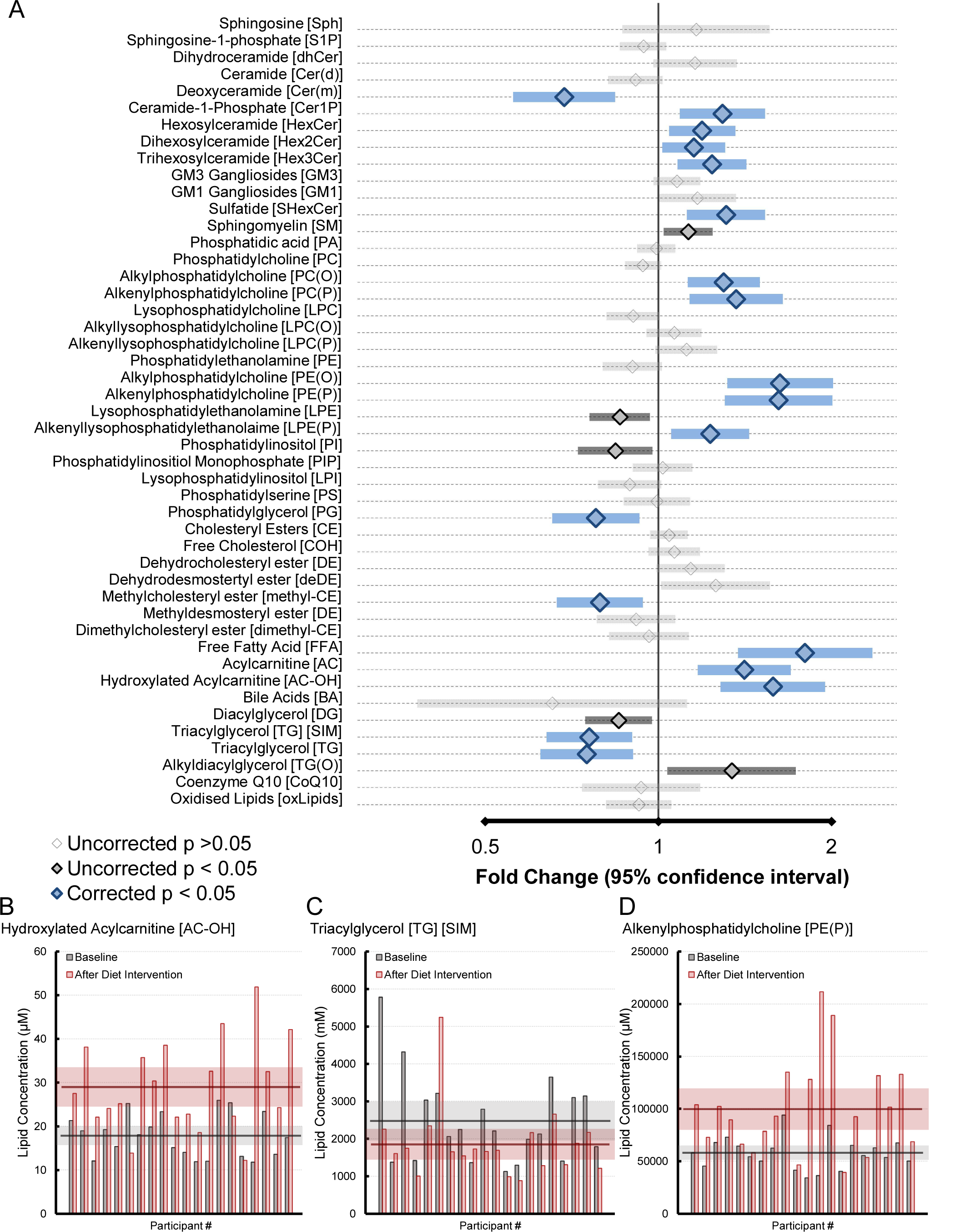
Lipid class associations with the modified Mediterranean ketogenic diet (MMKD). **A** Forest plot highlighting significant lipid classes associated with the 6-week MMKD (n = 20) from a linear mixed model. Units are in fold change of lipid concentration from baseline with 95% confidence intervals. Light grey, uncorrected p-value > 0.05, dark grey, uncorrected p-value < 0.05, blue, Benjamini-Hochberg corrected p-value < 0.05. **B-D** Relative concentration of each lipid class for each participant at baseline (grey) and follow-up (red). Horizontal bars represent mean concentration for baseline and follow up with 95% confidence intervals.

#### Fatty acid oxidation

Many of the changes associated with the plasma lipidome were expected with ketosis. The ketogenic diet would typically induce lipolysis, and ultimately shift towards fatty acid oxidation as an energy source rather than glucose.^42, 43^ We observed significant increases to the lipid classes acylcarnitines (AC, 1.41-fold increase, 1.20 - 1.67 95% CI, corrected p-value = 4.83 x 10_⁻³_), hydroxylated acylcarnitines (AC-OH, 1.58-fold increase, 1.31 - 1.91 95% CI, corrected p-value = 2.15 x 10^-3^) and free fatty acids (FFA, 1.80-fold increase, 1.40 - 2.31 95% CI, corrected p-value = 2.20 x 10^-3^) all indicative of increased lipid lipolysis and increased carnitine shuttling for fatty acid oxidation.^43^ These changes corresponded to declines in the triglyceride class (TG [NL], 0.75-fold change, 0.64 - 0.89 95% CI, corrected p-value = 1.20 x 10^-2^), thus together indicating conversion of more complex triglycerides into their free fatty acids and shuttling into mitochondria within cells and tissues for fatty acid oxidation.^44, 45^ Inspection of changes to lipid concentration at an individual level highlighted variable responses to MMKD for classes that increased (AC-OH, Figure 1B) and decreased (TG, Figure 1C).

#### Ether lipid metabolism

Unexpectedly, we observed a strong shift of ether glycerophospholipid concentrations within the peripheral lipidome after MMKD intervention. Ether lipids are a subgroup of lipids all sharing either the ether or vinyl-ether linkage in the sn1 position on the glycerol backbone.^46–48^ The strongest and most notable associations were observed with the two ethanolamine ether lipid classes, both in similar magnitudes, alkylphosphatidylethanolamine [PE(O)] concentration (1.63-fold change, 1.35 - 1.97 95% CI, FDR corrected p-value = 2.15 x 10^-3^) and the plasmalogen alkenylphosphatidylethanolamine [PE(P)] concentration (1.62-fold change, 1.33 - 1.97 95% CI, FDR corrected p-value = 2.15 x 10^-3^). In contrast, the non-ether glycerophospholipid, phosphatidylethanolamine (PE), was not associated with MMKD intervention (0.90-fold change, 0.80 - 1.01 95% CI, corrected p-value = 1.54 x 10^-1^) despite sharing many of the other structural characteristics to PE(O) and PE(P). Direct comparison of absolute lipid concentrations within individuals in response to the MMKD diet highlights differential responses (Figure 1D) with some individuals showing a 6-fold increase in PE(P) levels.

Similarly, the corresponding choline glycerophospholipid classes, alkylphosphatidylcholine, PC(O) and the plasmalogen alkenylphosphatidylcholine, PC(P), shared significant increases albeit with smaller effect sizes (PC(O) concentration, 1.30-fold increase, 1.15 - 1.47 95% CI, corrected p-value = 4.83 x 10^-3^ and PC(P) concentration, 1.37-fold increase, 1.16 - 1.61 95% CI, corrected p-value = 8.49 x 10^-3^, Figure 1A). These changes mirror the ethanolamine class with no associations observed in the more abundant phosphatidylcholine class (PC, 0.94-fold change, 0.87 - 1.01 95% CI, corrected p-value = 1.62 x 10^-1^). Interestingly, the atypical ether lipid, alkyldiacylglycerol [TG(O)] had nominally significant increases (1.34-fold increase, 1.04 - 1.73 95% CI, corrected p-value = 7.62 x 10^-2^, uncorrected p-value 3.73 x 10^-2^, Figure 1A) in stark contrast to the large reductions in circulating triglyceride content (Figure 1A). Despite sharing similar characteristics (3 long fatty chains on a glycerol backbone) these two lipid classes differ by the ether linkage on the sn1 position of the glycerol unique to ether lipids. Together, these appear to indicate shifts to ether lipid content in all pools with MMKD independent of bulk changes to glycerophospholipids.

#### Reductions in pathological sphingolipids

The accumulation of the unusual sphingolipid deoxyceramide (Cer(m)) has been associated with multiple metabolic disorders, more so than the well-reported ceramide class.^49, 50^ Deoxyceramides are synthesized via the promiscuous utilization of alanine or glycine instead of serine when synthesizing the sphingolipid backbone.^51^ We observed significant reductions in circulating deoxyceramides (0.69-fold change, 0.57 - 0.82 95% CI, corrected p-value = 4.83 x 10^-3^) in response to MMKD.

In general, many of the complex sphingolipid classes were increased significantly. As our lipidomic platform does not have the necessary isomeric separation to separate the structural isomers of the hexose group, more complex sphingolipids are annotated with the number of hexose groups rather than their explicit structure. All hexosylceramides were increased (hexosylceramide (HexCer), 1.19-fold change, 1.07 - 1.33 95% CI, corrected p-value = 2.00 x 10^-2^; dihexosylceramide (Hex2Cer), 1.15-fold change, 1.04 - 1.28 95% CI, corrected p-value = 3.13 x 10^-2^; and trihexosylceramide (Hex3Cer), 1.24-fold change, 1.10 - 1.40 95% CI, corrected p-value = 8.75 x 10^-3^). In human plasma, HexCer is predominately the glucosylceramide isoform (the other being the galactosylceramide), where Hex2Cer is the sphinglolipid with a lactose headgroup (lactosylceramide).

### Lipid species-specific responses to the modified Mediterranean ketogenic diet

While classes prove an overview of the changes at a higher level with dietary intervention, we observed intricate associations driven by specific species not seen at the class level. After FDR correction, we observed 393 lipid species (out of 784, Figure 2A) associated with MMKD intervention with 16 lipid species associated with the AHAD intervention (Supplementary Table 2).

**Figure 2:**
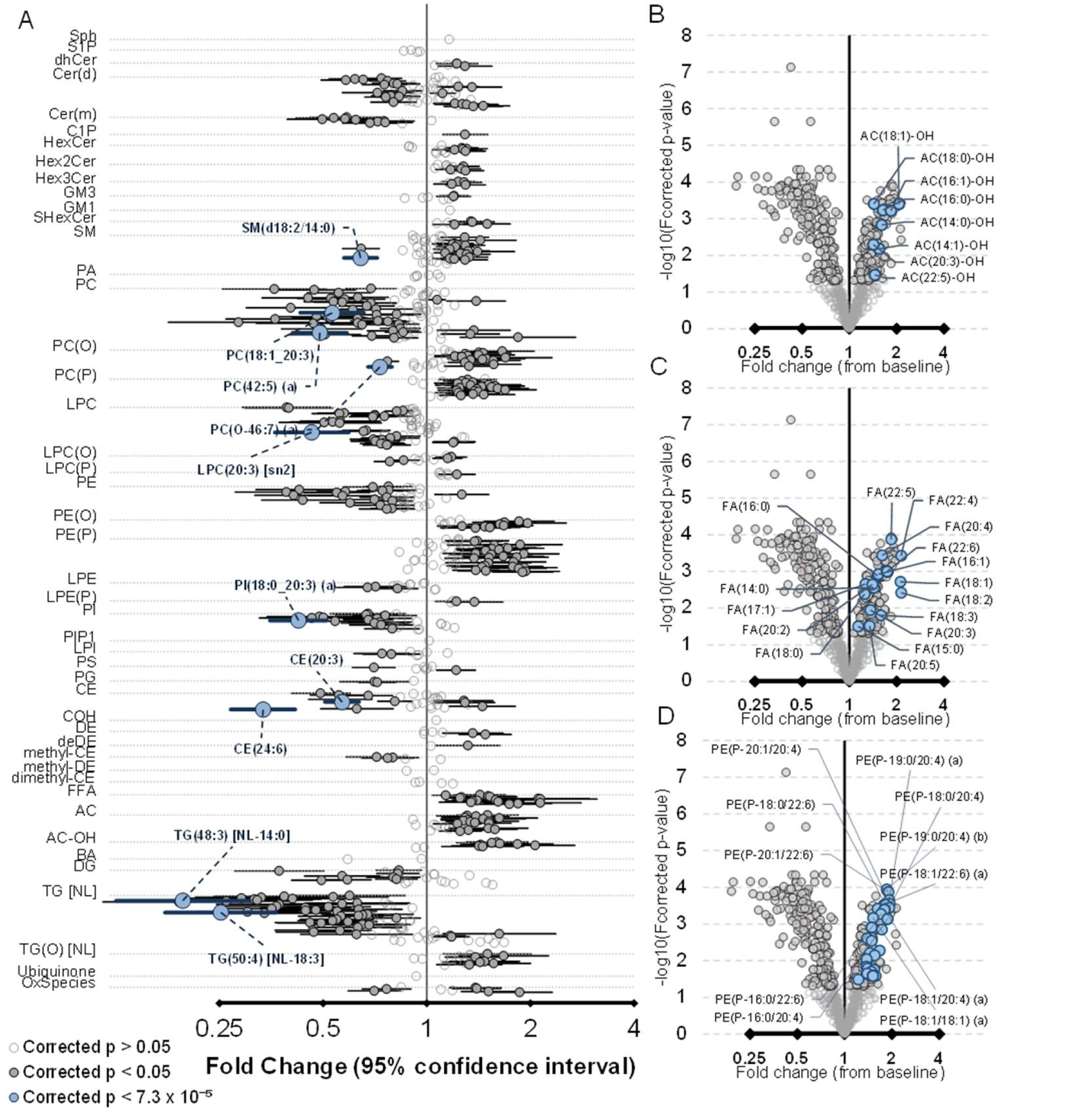
Lipid species associations with the modified Mediterranean ketogenic diet (MMKD). Associations of lipid species with 6-week MMKD (n = 20) obtained from a linear mixed model. Units are in fold change of lipid concentration from baseline with 95% confidence intervals. Light grey, corrected p-value > 0.05, dark grey, Benjamini-Hochberg corrected p-value < 0.05, blue, top 10 species ranked by p-value. **B-D-** Volcano plot highlighting specific lipid classes associated with MMKD intervention. *Abbreviations:* FA: Fatty Acid, NL: Neutral Loss, Ox: Oxidized. For the spelling of lipid species abbreviations, see Figure 1.

#### Fatty acid oxidation

Within the acylcarnitine and hydroxylated acylcarnitine classes, the associations with the largest effect size with MMKD intervention were the hydroxylated species with a 16:0, 16:1 and 18:1 fatty acid (AC(16:0)-OH, 1.63-fold change, 1.35 - 1.97 95% CI, corrected p-value = 5.84 x 10^-^□; AC(16:1)-OH, 1.84-fold change, 1.45 - 2.33 95% CI, corrected p-value = 6.06 x 10^-^□; AC(18:1)-OH, 2.07-fold change, 1.58 - 2.70 95% CI, corrected p-value = 3.93 x 10^-^□) (Figure 2B, Supplementary Table 2). Similar but smaller associations were seen in the non-hydroxylated species. All free fatty acid species were significantly increased with MMKD intervention, but the strongest associations were driven by several polyunsaturated fatty acids (PUFAs) (FA(22:5), 1.84-fold change, 1.47 - 2.31 95% CI, corrected p-value = 1.33 x 10^-^□; FA(20:4), 1.61-fold increase, 1.36 - 1.92 95% CI, corrected p-value = 3.68 x 10^-^□; FA(22:4), 2.14-fold increase, 1.62 - 2.81 95% CI, corrected p-value = 3.73 x 10^-^□; and FA(22:6), 1.74-fold increase, 1.38 - 2.19 95% CI, corrected p-value = 1.00 x 10^-3^) (Figure 2C, Supplementary Table 2). Declines to the triglyceride species were heavily skewed towards species with saturated or monounsaturated fatty acids, most notably species from 14:0, 16:0 and 16:1 classes (Supplementary Table 2), while species likely common in the diet (18:1, PUFA species) had minimal declines. Interestingly, several species with docosahexanoic acid (DHA, 22:6) had nominally significant increases with intervention in contrast to the class level changes, with a single species associated after FDR correction (TG(54:6) [NL-22:6], 1.63-fold increase, 1.12 - 2.37 95% CI, corrected p-value = 4.33 x 10^-2^). Importantly, we observed a significant decrease to CE(24:6) (0.33-fold change, 0.27 – 0.42 95% CI, corrected p-value = 2.27 x 10^-^□) with corresponding increases to CE(22:6) (1.45-fold change, 1.16 - 1.81 95% CI, corrected p-value = 1.18 x 10^-2^), which highlights peroxisome-driven β-oxidation for PUFA synthesis (Supplementary Table 2).

#### Ether lipid metabolism

While the abundant glycerophospholipids remain largely unaltered (e.g. PC(16:0_18:1), 0.98-fold change, 0.90 - 1.07 95% CI, corrected p-value = 7.08 x 10^-1^), significant species-level differences were seen, largely with PUFA fatty acids such as DHA increased (PC(16:0_22:6), 1.39-fold increase, 1.14 - 1.69 95% CI, corrected p-value = 1.16 x 10^-2^; PC(44:12), 1.84-fold increase, 1.25 - 2.71 95% CI, corrected p-value = 1.70 x 10^-2^). This is also reflected in the ethanolamine glycerophospholipids (Figure 2A, Supplementary Table 2).

Alterations to the ethanolamine ether lipids, both PE(O) and PE(P), were greater in highly PUFA species, including 20:4 (PE(P-18:0/20:4) concentration, 1.92-fold change, 1.52 - 2.42 95% CI, corrected p-value = 3.57 x 10^-^□) and 22:6 (PE(P-18:0/22:6) concentration, 1.83-fold change, 1.48 - 2.25 95% CI, corrected p-value = 2.98 x 10^-^□) (Figure 2D, Supplementary Table 2). The length of the sn1 position (alkyl/alkenyl) linkages did not appear to contribute differently to the association. Despite stronger changes with PUFAs in the sn2 position still strongly associated with the intervention (PE(P-18:0/18:1), 1.48-fold change, 1.25 - 1.76 95% CI, corrected p-value = 1.24 x 10^-3^) indicating an association driven by the ether lipids, not just increases to PUFAs. These effects are also reflected in the equivalent choline classes, PC(O) and PC(P).

### The impact of the MMKD intervention to the plasma lipidome was stronger in specific subgroups

#### Cognitive group interaction

Nine individuals were classified as MCI out of the 20 included as part of the trial. Interactions were weak, with 5 lipid classes presenting with a nominally significant (nominal p-value < 0.05) interaction. These classes are: phosphatidylethanolamine (PE), phosphatidylinositol (PI), phosphatidylglycerol (PG), ubiquinone (CoQ10) and triglycerides (Figure 3, Supplementary Table 3) in which the negative associations were stronger with the MCI group, notably the phosphatidylethanolamine (PE, 0.78-fold change, 0.67 - 0.91 95% CI, corrected p-value = 2.34 x 10^-2^) and triglyceride (TG, 0.62-fold change, 0.49 - 0.77 95% CI, corrected p-value = 6.61 x 10^-3^) (Figure 3, Supplementary Table 3) classes. Examination of the baseline concentration differences in these lipid classes highlighted nominally higher concentrations within the MCI group, including PE (1.50-fold difference, 1.04 - 2.17 95% CI, uncorrected p-value = 3.15 x 10^-2^) and TG (1.65-fold change, 1.07 - 2.54 95% CI, uncorrected p-value = 2.58 x 10^-2^) (Supplementary Table 3). Together, this indicates a small normalizing effect of the perturbed lipidome identified in the MCI group of this smaller study. This finding requires further investigation in studies designed with sufficient statistical power.

**Figure 3:**
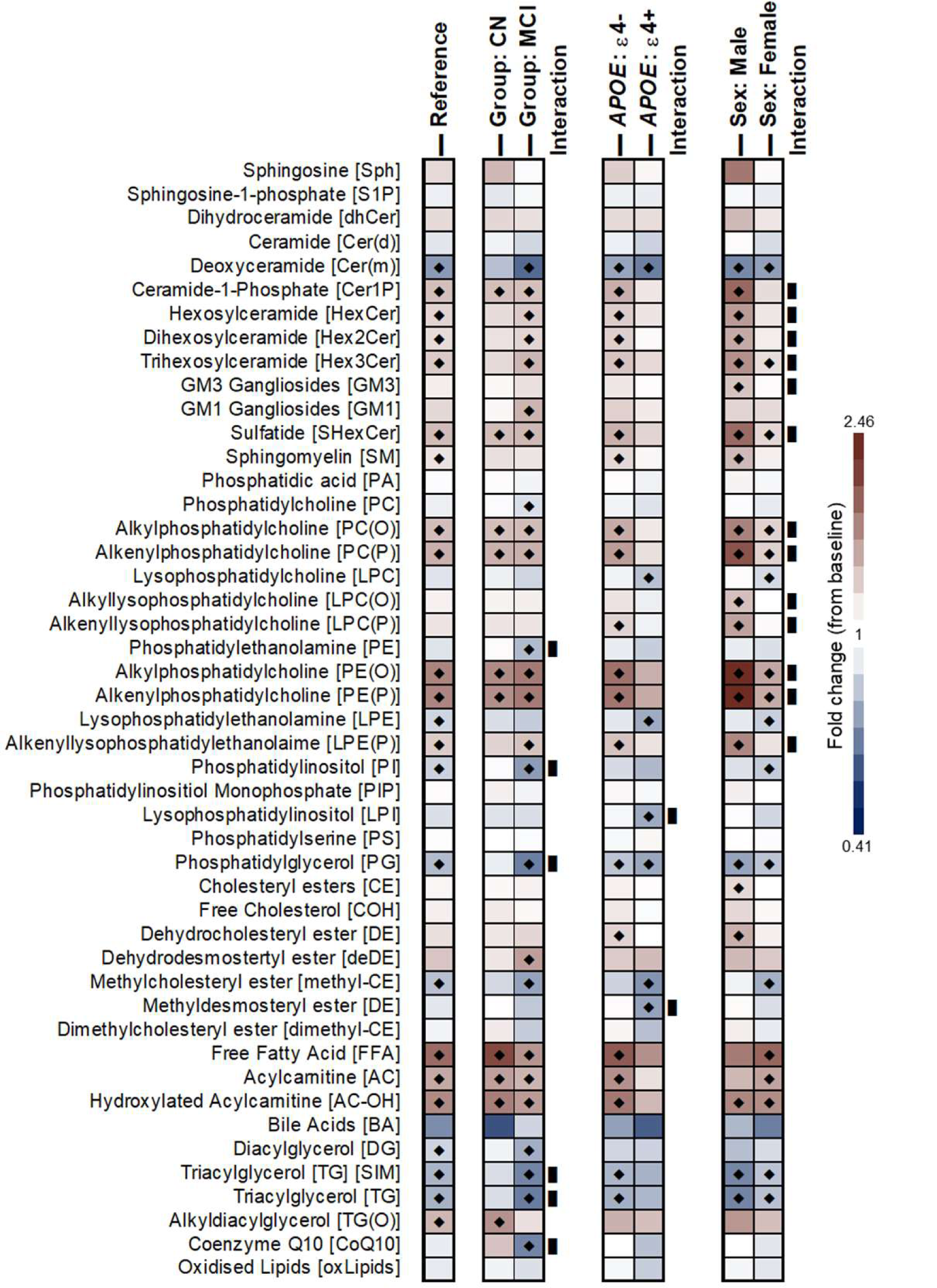
Differential plasma lipid response with the MMKD intervention based on subgroup interactions. A heatmap highlighting lipid associations with MMKD intervention stratified using an interaction term for cognitive group (**left**), *APOE* genotype (**middle**) and sex (**right**). Reference denotes the unadjusted linear mixed model on lipid associations with MMKD intervention. Black diamonds, nominal p-value < 0.05, black block, nominal interaction p-value < 0.05. Both nominal and corrected p-values are in Supplementary Tables 3-5. *Abbreviations:* APOE: Apolipoprotein E, CN: Cognitively Normal, MCI: Mild Cognitive Impairment, MMKD: Modified Mediterranean-Ketogenic Diet.

#### APOE genotype interaction

Six participants had a copy of the *APOE* □4 allele. Overall, we observed only minor interactions between lipid concentrations after MMKD by *APOE* □4 status (Supplementary Table 4). However, several plasma lipid changes differed by APOE status after MMKD intervention, in which lipid associations were only significant for the non-APOE □4 individuals; for example: PE(O) (*APOE* □4 – PE(O), 1.37-fold change, 0.97 - 1.93 95% CI, corrected p-value = 3.35 x 10^-1^ and non-*APOE* □4 – PE(O), 1.76-fold change, 1.40 - 2.20 95% CI, corrected p-value = 2.34 x 10^-3^), in which lipid associations were only significant for the non-*APOE* □4 individuals. This effect was observed for all impacted lipid classes (Supplementary Table 4). While the reduction in association can’t be completely attributed to the *APOE* □4 genotype in this study, it’s an interesting observation that warrants further investigation.

#### Sex specific interactions

We observed differences in the strength of associations with multiple ether lipid classes when examining sex-specific interactions. Despite the difference in numbers (5 males, 15 females), the associations were stronger in males (PE(P), 2.52-fold change, 1.82 - 3.48 95% CI, corrected p-value = 9.37 x 10^-^□) compared to females (PE(P), 1.40-fold change, 1.16 - 1.68 95% CI,corrected p-value = 2.45 x 10^-2^) with a nominally significant interaction p-value (p = 6.48 x 10^-3^) (Figure 3, Supplementary Table 5). This difference was not attributed to specific lipid class concentrations between males and females (Supplementary Table 5).

### Alteration to the plasma lipidome with the Modified Mediterranean Ketogenic Diet is inversely related to the signature observed in Alzheimer’s disease

One of the striking changes associated with the MMKD was its impact on several lipid classes previously identified to associate with AD.^34^ We correlated the lipid class associations identified with MMKD intervention to associations previously reported for prevalent and incident AD in The Australian Imaging, Biomarker & Lifestyle Flagship Study of Ageing (AIBL) and ADNI studies. The distinct AD lipidome signature is highlighted by alterations in sphingolipid: dihydroceramides (dhCer), Hex3Cer, GM3 gangliosides (GM3), GM1 gangliosides (GM1), sulfatides, and ether lipids PC(O), PC(P), PE(O), PE(P), TG(O)^34^. An inverse relationship was observed for most of the classes, notably for all of the ether lipid classes: PE(P), PE(O), PC(O), PC(P), and TG(O) for both prevalent and incident disease (Figure 4A and B, Supplementary Table 6). It was noted that some of the glycosphingolipids (HexCer and Hex2Cer) were positively associated with AD risk but were increased with MMKD intervention (Figure 4A).

**Figure 4:**
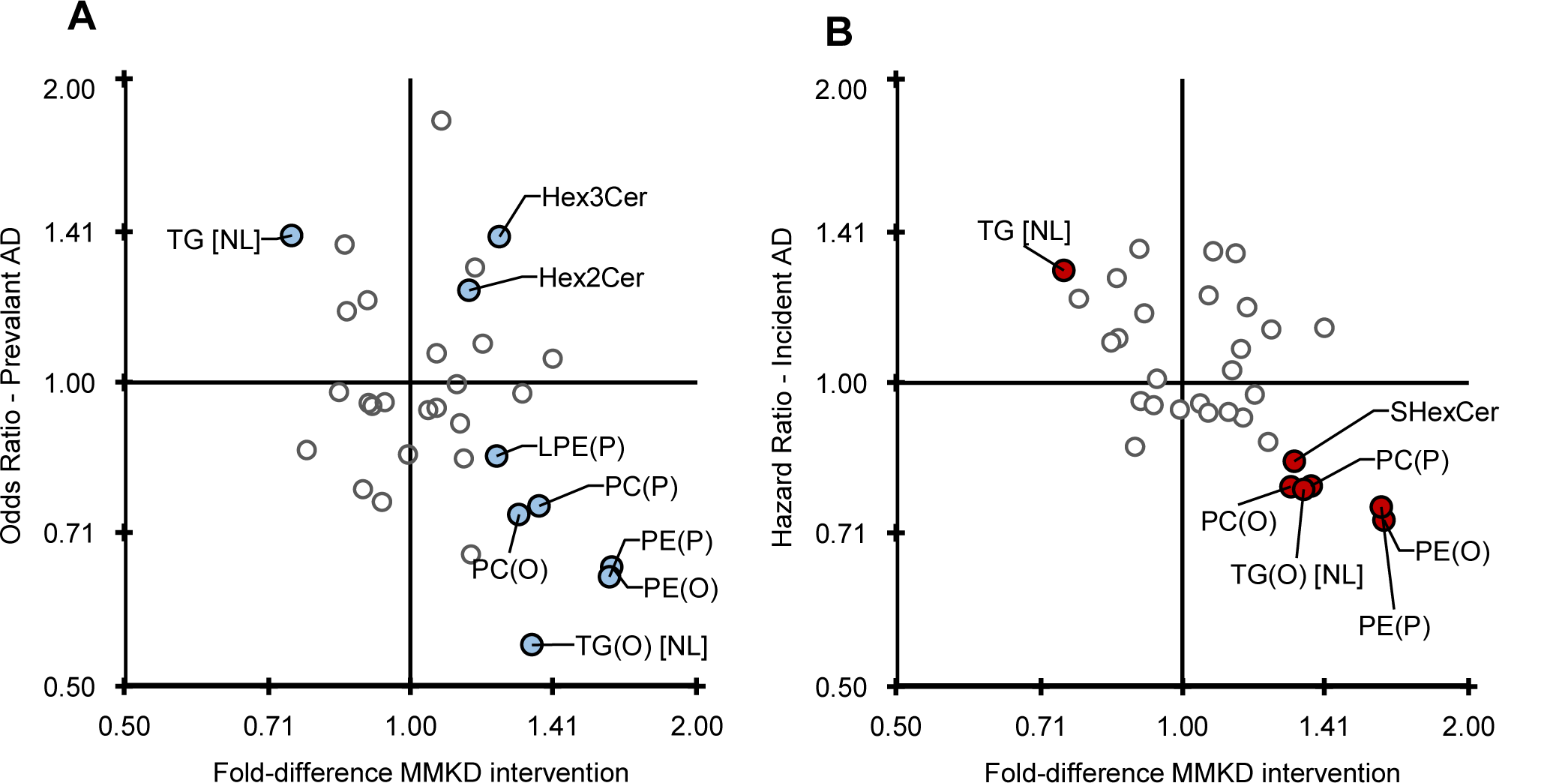
Inverse relationship between plasma lipid changes with MMKD intervention and signatures associated with Alzheimer’s disease. A scatter plot correlating lipid class associations with MMKD intervention and Alzheimer’s disease (data previously reported Huynh et al. 2020). **A** - Lipid classes nominally associated with both MMKD and prevalent/existing AD (highlighted in blue, class totals, meta-analysis of both AIBL and ADNI) and **B** – Lipid classes nominally associated with both MMKD and incident/future risk of AD (highlighted in red). *Abbreviations:* AD: Alzheimer’s Disease, Hex2Cer: Dihexosylceramide, Hex3Cer: Trihexosylceramide, LPE: Lysophosphatidylethanolamine, MMKD: Modified Mediterranean-Ketogenic Diet, NL: Neutral Loss, PC: Phosphatidylcholine, PE: Phosphatidylethanolamine, SHexCer: Sulfatide, TG: Triglyceride.

### Changes to several CSF and cognitive markers of Alzheimer’s disease correlated with the changes in the peripheral lipidome

We next examined whether any of the lipid changes identified in plasma after diet intervention were associated with changes observed in various AD blood, CSF and cognition measures. Significant positive correlations in changes to both lipid classes and blood biochemical measurements were observed for all clinical measures (total cholesterol, triglycerides, high-density lipoprotein cholesterol, low-density lipoprotein cholesterol and very low-density lipoprotein cholesterol) as expected (Figure 5). Species-dependent correlations were observed in which clinical measurement of triglycerides were not associated with species esterified with a 22:6 (DHA) fatty acid (Figure 6). Correlations between the changes in lipid classes and fasting ketones level were predominately seen in the free fatty acid lipid class (FFA, r = 0.62) and were inversely correlated with total sphingosine 1 phosphate (S1P, r = −0.54, Figure 5).

**Figure 5:**
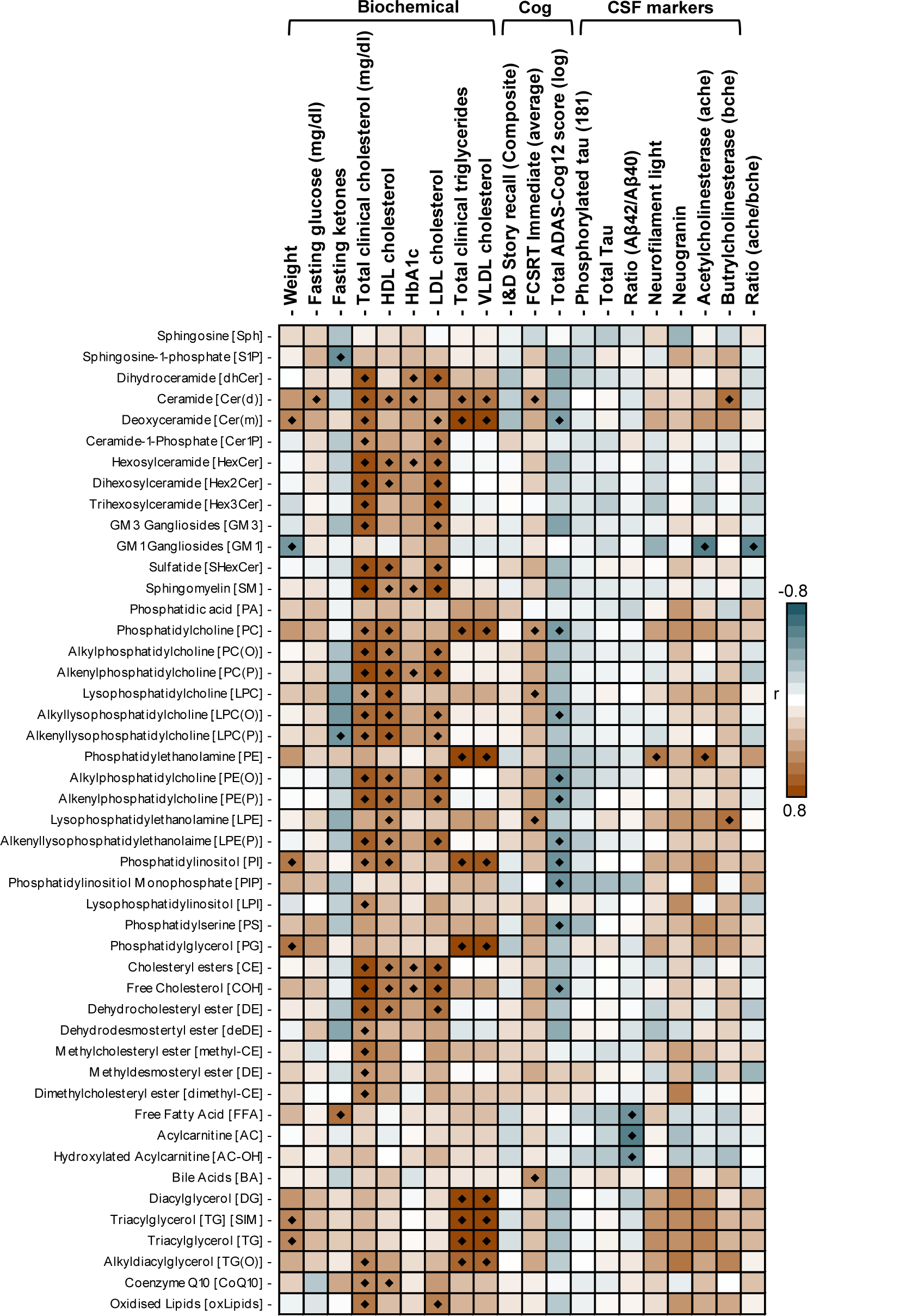
Correlated changes between plasma lipid totals (MMKD intervention) with Alzheimer’s disease risk factors, markers and outcomes. A Pearson correlation between changes in the total of each lipid class with changes in different biochemical, cognition and CSF markers. Black diamond, p < 0.05. *Abbreviations:* ADAS-COG12: Alzheimer’s Disease Assessment Scale-Cognitive, Cog: Cognition, CSF: Cerebrospinal Fluid, FCSRT: Free and Cued Selective Reminding Test, HbA1C: Hemoglobin A1c, HDL: High-Density Lipoprotein, I&D Story Recall: Immediate and Delayed Story Recall composite, LDL: Low-Density Lipoprotein, MMKD: Modified Mediterranean-Ketogenic Diet, VLDL: Very-Low-Density Lipoprotein.

**Figure 6:**
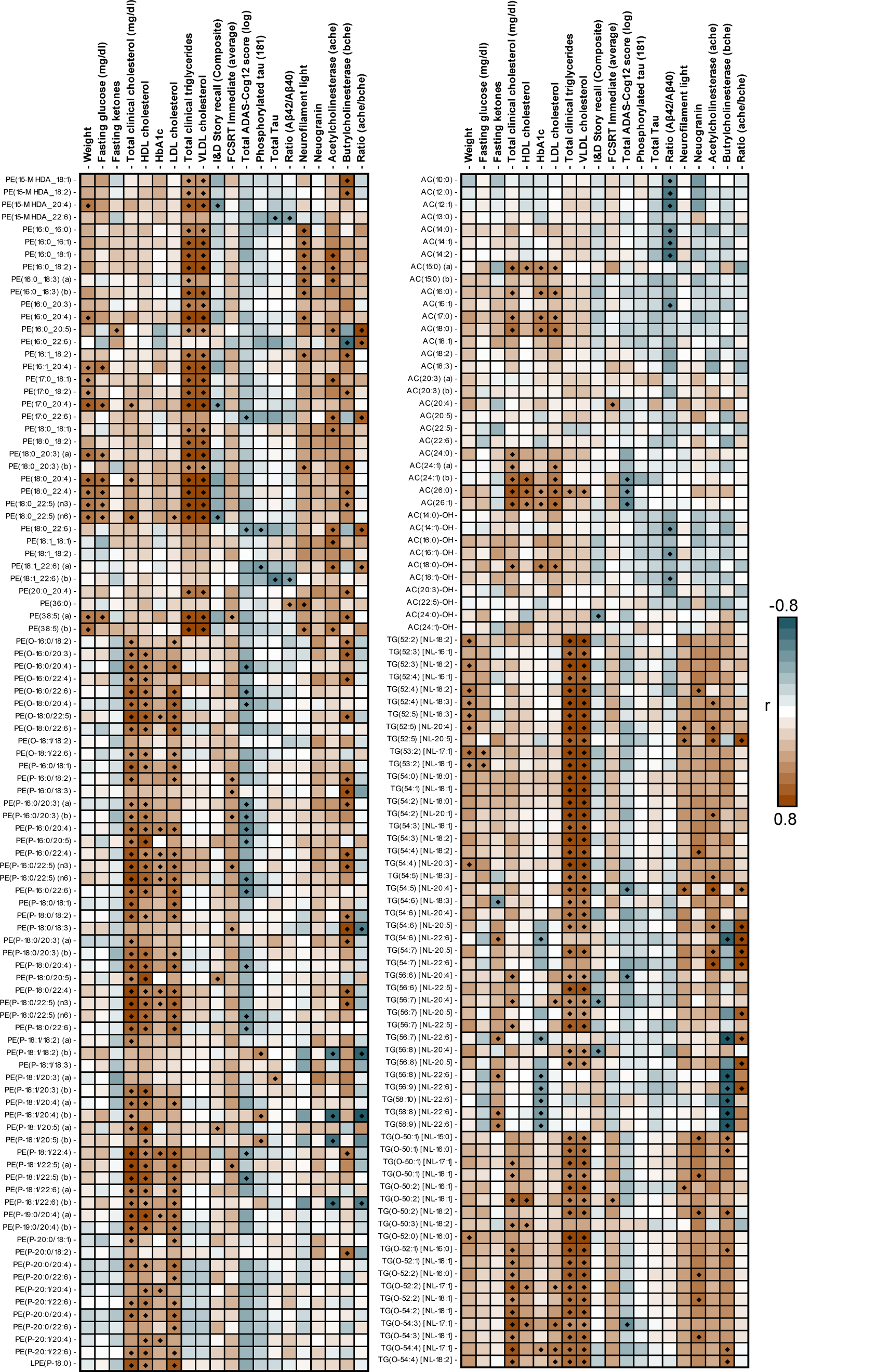
Correlated changes between plasma lipid species (MMKD intervention) with Alzheimer’s disease risk factors, markers and outcomes. Pearson correlation between changes in select lipid species with changes in different biochemical, cognition and CSF markers. Black diamond, p < 0.05. *Abbreviations:* AC: Acylcarnitine, ADAS-COG12: Alzheimer’s Disease Assessment Scale-Cognitive, CSF: Cerebrospinal Fluid, FCSRT: Free and Cued Selective Reminding Test, HbA1C: Hemoglobin A1c, HDL: High-Density Lipoprotein, I&D Story Recall: Immediate and Delayed Story Recall composite, LDL: Low-Density Lipoprotein, MMKD: Modified Mediterranean-Ketogenic Diet, TG: Triglyceride, PE: Phosphatidylethanolamine, VLDL: Very-Low-Density Lipoprotein.

Changes to ADAS-Cog12 (log-transformed) was inversely correlated with changes to many ether lipid classes (PE(O), r = −0.52; PE(P), r = −0.49) (Figure 5) with stronger species-specific correlations observed in PUFA plasmalogen species (PE(P-16:0/22:6), r = −0.57; PE(P-18:0/22:6) (n6), r = −0.54) (Figure 6). With MMKD intervention, inverse correlations were observed in changes to very long chain acylcarnitines (AC(26:0), r = −0.50; AC(26:1), −0.57) but not other acylcarnitine species (Figure 6, Supplementary Table 7). In contrast to ADAS-Cog12, there were fewer correlations observed as significant with changes to the other cognitive measures. FCSRT Immediate (average) had positive correlations with changes seen in many omega-6 esterified species (LPE(20:4) [sn1], r = 0.69, AC(20:4), r = 0.52) while Immediate and delayed story recall composite (I&D Story recall) was positively correlated with some changes in 20:5 species (PE(P-18:0/20:5), r = 0.50) (Supplementary Table 6).

The changes to several CSF biomarkers for AD were correlated with lipid changes after MMKD intervention (Figure 5, Supplementary Table 7). MMKD-associated changes in the ratio of Aβ42/40 were inversely correlated with changes to total free fatty acids and acylcarnitines (Figure 5). Unlike associations with ADAS-Cog12, the stronger acylcarnitine correlations were predominately medium in length and in species implicated in the fatty oxidation steps (AC(14:2), r = −0.67; AC(14:1)-OH, r = −0.54) (Figure 6). While both total and phosphorylated tau changes were not correlated with class total changes, species-specific inverse relationships were observed with 22:6 containing phospholipids (Supplementary Table 7). Interestingly, four subspecies of plasmalogens with an 18:1 (b) isoform on the vinyl-ether were positively associated with changes in phosphorylated tau (Supplementary Table 7). The 18:1 (b) isoform corresponds to the more abundant 18:1 omega-9 isoform. While changes to neurofilament light were weakly inversely related to plasmalogens, they were strongly correlated with changes to saturated and monounsaturated PE species (Figure 6). Similar to phosphorylated tau, an isoform specific correlation between plasmalogen and the ratio of acetylcholinesterase / butyrylcholinesterase was observe. This effect was predominately driven by the 18:1 (b) isoform on the vinyl ether (PE(P-18:1/20:4) (b), r = −0.80) (Figure 6, Supplementary Table 7) with significant positive correlations with omega-3 triglycerides (TG(54:7) [NL-22:6], r = 0.91) (Figure 6).

## DISCUSSION

This is the first clinical study to assess the impact of a ketogenic dietary intervention on the lipidome in patients at risk for AD. We used a powerful targeted lipidomics platform to map the effect of a KD on the plasma lipidome to better define the metabolic changes that underlie the potential therapeutic benefits in AD. We found four key findings: (1) MMKD but not AHAD had a large impact on the plasma lipidome, (2) some lipidomic changes were more pronounced in those with baseline cognitive impairment, and most importantly (3) MMKD-induced changes were inversely related to patterns previously found in AD; which suggests that (4) MMKD may be effective in reversing the undesirable AD lipidomic profile.

We previously showed that a modified ketogenic diet (MMKD) improved key CSF AD biomarkers, imaging measures, and cognition in a pilot study of adults at risk for AD.^52^ In the present study, we sought to examine the effects of this intervention on the plasma lipidome. The MMKD had a considerable impact on the plasma lipidome at both the class and species level, although we found strong MMKD-associated changes in lipid species that were not seen at the class level.

### Impact on fatty acid oxidation and metabolism

Ketosis is the process in which the body shifts to metabolize fats rather than carbohydrates and is observed in low glucose situations. This process is essential for providing peripheral ketones to the brain for energy, as the brain is unable to metabolize fats in the same manner. Our data reflects this process in the periphery, where complex triglycerides are broken down, resulting in the corresponding increases to free-or carnitine-bound fatty acids. In ageing and AD, brain energy homeostasis plays an important role and is believed to be impaired, with some research indicating a central dysregulation of glucose metabolism.^53, 54^ Ketone bodies transported into the brain are believed to provide an alternative energy source to rectify some of the perturbation in glucose metabolism in AD.^55^

All free fatty acid species were increased after the MMKD, but the strongest associations were driven by several PUFAs (22:5, 22:4 and 22:6). This is similarly reflected in the minimal declines of only PUFA esterified triglycerides. Both hydroxylated species of acylcarnitines (16:0, 16:1 and 18:1 fatty acid) increased, with similar associations in the non-hydroxylated species, which highlights both increases to carnitine shuttling, and hydroxylation for oxidation. Both TG and FFA can cross the blood-brain barrier, albeit at different rates and through different mechanisms.^56^ A reduction in triglycerides is an established effect of low fat, ketogenic diets, and is thought to confer beneficial effects.^57–59^ Our results indicate that MMKD could impact disease-related pathways by increasing circulating ketone bodies and increasing the ratio of circulatory free fatty acids and acylcarnitines to triglycerides, while also significantly enriching favorable PUFAs (including 22:6).

### Impact on ether lipid metabolism and peroxisomal activity

Dysregulation of ether lipid metabolism has been linked to neurodegenerative disease, metabolic disorders, and cancer.^46^ Plasmalogens, specifically, are lower in the blood and brains of those with Alzheimer’s and related to worse clinical outcomes (ADAS-Cog).^20, 26, 29, 60, 61^ Indeed, preclinical studies support interventions aimed at increasing plasmalogens, which may reduce neuroinflammation and neurodegeneration, mediated in part through enhanced AKT and ERK signaling.^62, 63^ We previously highlighted that the plasma level of ether lipids, notably alkyldiacylglycerols and TG(O), mediate nearly one-third of the protective effect of the *APOE* □2 allele in AD.^41^ Thus interventions aimed at increasing TG(O), such the MMKD, may either provide beneficial effects in patients without *APOE* □2 or further potentiate such effects in those with the *APOE* □2 allele.

We unexpectedly found an increase in several ether lipid classes after the MMKD intervention, including plasmalogens (PE(P) and PC(P)), choline and ethanolamine alkyl-ether lipids (PC(O and PE(O)), and the ether glycerolipids (TG(O)). To date, no dietary intervention has demonstrated systemic alterations to these lipid species and classes to such a degree. While both PE(O) and PE(P) increased after the MMKD, the structurally similar non-ether glycerophospholipid phosphatidylethanolamine (PE) did not change. This stark contrast was also observed within the TG/TG(O) classes, with prominent increases to TG(O) despite decreases to TG. Together, these results suggest that the MMKD may selectively impact the ether lipid classes rather than bulk changes to the lipidome as a whole.

Ether lipids are unique in their synthetic pathway and require specific enzymes found only in the peroxisome.^46^ Plasmalogens have an additional step that involves plasmanylethanolamine desaturase 1 (PEDS1) that introduces a double bond on the alkyl-linkage, which results in the vinyl-ether bond unique to this lipid class.^64^ Few approaches have been identified to increase ether lipid content in human plasma to such an extent. Alkylglycerols and similarly structured lipid metabolites can increase circulating ether lipid levels by acting as direct precursors that bypass the rate limiting peroxisomal step^65^ and are found in modest levels in several marine oils including squid and shark liver oil.^65^

Our results together indicate a significant upregulation of ether lipid synthesis, likely driven by increased peroxisomal number or function.^46^ It is unlikely that that lipid signature of the changes with the MMKD are simply attributed to the increased dietary intake of specific food groups. Indeed, aside from increases to ether lipids as a whole, this effect is further corroborated with significant decreases to CE(24:6) with corresponding increases to CE(22:6) (Supplementary Table 2). Peroxisomal very long chain fatty acid β-oxidation is required for *de novo* synthesis of 22:6 from other precursors.^66^ Similarly, one existing study indicated potential improvements to peroxisomal substrate oxidative capacity in skeletal muscle with ketogenic diet and exercise.^67^ As plasmalogens and ether lipids are rate limited in their synthesis within the peroxisome, our results indicate that MMKD impacts these metabolic pathways.

### Impact on sphingolipids

The MMKD-associated reduction in deoxyceramides may also be beneficial. Increased deoxyceramides are an established feature of metabolic disorders and have been associated with obesity in patients with type 2 diabetes mellitus.^49^ Understanding the impact of deoxyceramides in metabolic dysfunction is important given the strong relationship between systemic metabolic conditions and AD.^68^ Deoxyceramides may also contribute to multi-system age-related changes. An elegant study that compared multiple tissues (brain, fat, muscle, liver) in young versus older mice showed that deoxyceramides, as well as deoxydihydroceramides and ether-linked diacylglycerols, were increased in aging and possibly related to senescence.^69^ While we found no significant associations with ceramides when examined as a lipid class (Figure 1A), we found species-specific relationships driven by the length of the sphingoid base. Species with a d16:1 sphingoid base were reduced, and species with more abundant d18:1 sphingoid bases were increased after the MMKD. This further supports for the need to examine species-level effects.

### Interaction with clinical subgroups and variable response

One of our most striking findings was that the impact of the MMKD on the plasma lipidome appears dependent on cognitive group. While many of the lipid classes changed independently of baseline cognitive diagnosis, several lipid changes were more prominent in participants with MCI relative to those who were cognitively normal. The MCI group particularly had larger decreases to triglycerides and phosphatidylethanolamine, representing a favourable lipid profile.^34, 70–72^ Similarly, individuals with the *APOE* □4 genotype were less impacted by the MMKD, which highlights the potential for the *APOE* genotype to influence lipid metabolism. Little is known about how cognitive status and *APOE* genotype impacts systemic lipid metabolism and response to dietary intervention, thus this work provides a valuable foundation for future studies. While most patients had a similar pattern of lipidome changes in response to the MMKD (Figure 1B-D), there was marked variation in the lipidome alteration among individuals. A subset of participants displayed a substantial response to the diet, with some experiencing a nearly six-fold increase in lipid classes from baseline after the 6-week intervention. This highlights the importance of identifying those “super-responders” who may exhibit a heightened response to the diet, as it could facilitate the development of personalized therapies for those at risk of AD.

### Potential restoration of dysregulated lipid Alzheimer’s disease signature

The MMKD altered the lipidome in a manner that was opposite from the lipid profiles observed in both individuals with AD cross sectionally (Figure 4A) and development longitudinally (Figure 4B) from two independent studies of AD.^40^ In subsequent exploratory analyses, we found that the corresponding increases of PE(P) in circulation from the MMKD were associated with decreased ADAS-Cog12 score (Supplementary Figure 3). This suggests that participants with a greater increase in plasmalogens, a lipid class reduced in AD,^60, 63^ had more benefit on a clinically meaningful endpoint.

Importantly, our results suggest that the pathologic AD lipidome signature can be attenuated through a dietary intervention. Modulating the peripheral lipidome with specific dietary intervention is far less invasive, less costly, and less prone to adverse effects than upcoming monoclonal antibody treatments,^73^ making it far more suited for early intervention considering the risk-benefit if efficacy can be demonstrated. Utilization of a dietary intervention like the MMKD as a supplementary intervention (combination therapy similar to the FINGER trial^74^ or with monoclonal antibody therapy) could provide tangible clinical benefits.

### Strengths and Limitations

The present study features several strengths, including its prospective crossover design, advanced lipidomic methods, and well-characterized data sample. Specifically, the study design enables a more accurate evaluation of the effects of a ketogenic (and low-fat) diet on lipidomics in AD by having each participant serve as their own metabolic control. This approach effectively controls other patient characteristics, making it more likely to detect diet-related changes. Furthermore, it is likely that the benefits of a ketogenic dietary intervention extend beyond ketosis alone and supports the use of a comprehensive diet aimed at inducing ketosis rather than solely supplementing ketone bodies or medium chain triglycerides as an attempt to achieve similar efficacy.^58, 59, 75–79.^

There are several limitations that must be acknowledged. The sample size is small, which limits the generalizability of our findings. While we observed robust diet-associated lipid changes, these findings should be interpreted within the context of the sample size. It is also challenging to adequately assess the effects of *APOE* genotype, which plays a crucial role in lipid metabolism and AD.

### Next steps

We observed a profound alteration in plasma lipids following implementation of the MMKD. However, it remains uncertain as to whether these changes are primarily driven by the metabolic effects of ketosis, by the specific constituents of the MMKD, or the combination of both. It is noteworthy that both cognitive groups were prescribed the same target diet, with the same dietician and menu, and yet the MCI group displayed a more pronounced response. Future investigations will aim to gain a deeper understanding of the underlying mechanisms. Whether the MMKD-induced modifications in the plasma lipidome are associated with beneficial brain changes will be further examined using the novel blood-based biomarkers for Alzheimer-related processes.

## Conclusion

This study represents the first clinical investigation to examine the effects of a modified ketogenic dietary intervention on the lipidome in adults at risk for AD. Our findings demonstrate that a modified ketogenic diet led to considerable modifications in the plasma lipidome. The observed changes were generally beneficial, and more pronounced in participants with cognitive impairment. Notably, the MMKD-associated lipid changes were inverse to an established AD lipidome signature. These results suggest that a modified ketogenic diet may serve as a therapeutic intervention to reverse pathologic AD lipid changes, but more extensive mechanistic studies are required. The lower relative cost and risks associated with an MMKD enhances the appeal of this approach for use in prevention or in combination therapy for early symptomatic AD.

## ONLINE METHODS

### Study Participants

We included participants at risk for AD based on baseline cognitive dysfunction (MCI) diagnosed by expert physicians and neuropsychologists using National Institutes of Health - Alzheimer’s Association MCI criteria,^80^ or who had subjective memory complaints using the Alzheimer’s Disease Neuroimaging Initiative (ADNI) criteria.^81^ All had prediabetes as defined by American Diabetes Association guidelines, with a screening hemoglobin A1c of 5.7-6.4%.^82^ Study exclusion criteria included a prior diagnosis of neurological or neurodegenerative illness (except MCI), major psychiatric disorder (well-controlled depression was allowed), prior stroke, current use of diabetes and lipid lowering medications, or current use of medications that impact central nervous system activity (i.e., anti-seizure medications, anti-psychotics, opioids).

The protocol was approved by the Wake Forest Institutional Review Board (ClinicalTrials.gov Identifier: NCT02984540), and written informed consent was obtained from all participants and/or their study partners. Participants were medically supervised by clinicians, with safety monitoring overseen by the Wake Forest Institutional Data and Safety Monitoring Committee.

### Procedure

This was a randomized crossover pilot trial in which participants consumed either a Modified Mediterranean-Ketogenic Diet (MMKD) or the American Heart Association Diet (AHAD) for 6 weeks. This was followed by a 6-week washout period, after which the diet not previously used was consumed for 6 weeks (Supplementary Figure 1). Participants were instructed to resume their pre-study diet during the washout period. Randomization to the initial diet was through a random number generator. See Neth et al.^52^ for complete details.

### Diet Intervention and Education

The experimental diet (MMKD) was a modified ketogenic diet, which has increasingly been utilized in medically intractable epilepsy due to its increased tolerability and similar efficacy to the traditional ketogenic diet.^13^ The target macronutrient composition (expressed as % of total calories) was 5-10% carbohydrate, 60-65% fat, and 30% protein. Note that this is in line with the “Modified Atkins Diet”, although it contains less fat than a traditional KD, which contains about 90% fat by calories.^13^ Daily carbohydrate consumption was targeted at <20g/day. Participants were encouraged to avoid store-bought products marketed as “low-carbohydrate” and artificially sweetened beverages. Extra virgin olive oil was supplemented, and participants were encouraged to eat fish, lean meats, and nutrient rich foods as the source of carbohydrates (i.e., green leafy vegetables, nuts, berries).

Our control diet (AHAD) was adapted from the low-fat American Heart Association Diet.^83^ The target composition of the AHAD was 55-65% carbohydrates, 15-20% fat, and 20-30% protein. Daily fat intake was targeted at <40g/day. Participants were encouraged to eat plentiful fruits, vegetables, and fiber-laden carbohydrates.

A registered dietitian developed daily meal plans for each participant based upon their food preferences and caloric needs as determined by a pre-study 3-day food diary, body composition, and activity level. Participants had weekly diet education visits (either in-person or by phone) starting one week prior to the start of each diet and continuing throughout the remainder of the intervention. Participants maintained a daily food record that was reviewed at these visits. Both diets were eucaloric and targeted to each participant’s baseline caloric needs with a goal of keeping weight neutral throughout the course of the study. Participants were asked to keep their exercise and physical activity level stable throughout the study. Adherence was assessed by capillary ketone body (beta-hydroxybutyrate) using the Nova Max Plus® (http://www.novacares.com/nova-max-plus/) and using participant subjective reports. Participants were required to supply their own food with a food stipend of $25/week provided to defray higher food costs. Participants received a daily multivitamin supplement (Centrum® Silver®) during both diets. The following supplements were not allowed for the duration of the study: resveratrol, CoQ10 (coenzyme Q10), coconut oil/other medium chain triglyceride-containing supplements, or curcumin.

### Blood Collection and Processing

Fasting blood was collected before and after each diet, and at follow-up. Samples were immediately placed on ice and spun within 30 minutes at 2200 rpm in a cold centrifuge for 15 minutes. Plasma, serum, and red blood cells were aliquoted into separate storage tubes and flash frozen at −80°C until analyzed.

### Lipid extraction and lipidomic analysis (Plasma)

Serum samples (10 µL) were extracted using a single-phase process comprised of 90 µl of butanol:methanol 1:1 and 10 µL of an internal standard mix containing a mix of non-physiological and stable isotope-labelled lipid standards (Supplementary Table 8). In brief, samples were mixed with the extraction solvent, vortexed and sonicated on a sonicator bath for 1 hour. They were subsequently centrifuged, and the supernatant transferred into glass vials with inserts for mass spectrometry analysis.

Lipidomics was performed as described previously with modifications.^84^ Analysis of plasma extracts was performed on an Agilent 6495C QQQ mass spectrometer with an Agilent 1290 series HPLC system and a single ZORBAX eclipse plus C18 column (2.1 x 100mm 1.8µm, Agilent) with the thermostat set at 45°C. Mass spectrometry analysis was performed in both positive and negative ion mode with dynamic scheduled multiple reaction monitoring (Supplementary Table 8). More details about our latest methodology to generate these lipids are kept up to date on our lab website (https://metabolomics.baker.edu.au/method/). The running solvent consisted of solvent (A) 50% H2O / 30% acetonitrile / 20% isopropanol (v/v/v) containing 10mM ammonium formate and 5µM medronic acid, and solvent B: 1% H2O / 9% acetonitrile / 90% isopropanol (v/v/v) containing 10mM ammonium formate.

The following mass spectrometer conditions were used: gas temperature 150°C, gas flow rate 17L/min, nebulizer 20psi, Sheath gas temperature 200°C, capillary voltage 3500V, and sheath gas flow 10L/min. Isolation widths for Q1 and Q3 were set to “unit” resolution (0.7 amu).

For the chromatography, we used a stepped linear gradient with a 16-minute cycle time per sample and a 1 µL sample injection. The sample analytical gradient was as follows: starting with a flow rate of 0.4 mL/minute at 15% B and increasing to 50% B over 2.5 minutes, then to 57% over 0.1 minutes, to 70% over 6.4 minutes, to 93% over 0.1 minute, to 96% over 1.9 minutes and ramped to 100% over 0.1 minute. The solvent was then held at 100% B for 0.9 minutes (total 12.0 minutes). Equilibration was started as follows: solvent was decreased from 100% B to 15% B over 0.2 minutes and held for a final run time of 16 minutes.

To align the results to any future datasets, the NIST 1950 SRM sample (Sigma) was included as a reference plasma sample at a rate of 1 per 40 samples. As indicators of between-batch quality, pooled QC samples and technical QC samples were injected at a rate of 1 per 20 samples. Relative quantification of lipid species was determined by comparison to the relevant internal standard. Several lipid classes do not have direct internal standards and the closest structural lipid standard was used (Supplementary Table 8). Lipid class total concentrations were calculated as the sum of individual lipid species concentrations (detailed in Supplementary Table 8).

### Lumbar Puncture

Participants underwent a lumbar puncture for CSF collection after a 12-hour fast at baseline and after each diet. Participants were placed in the seated or lateral decubitus position per study clinician preference. Using a 25-gauge needle, the L3-4 or L4-5 interspace was infiltrated with 1% lidocaine for local anesthesia. Using a 22-gauge Sprotte needle to drip, up to 25ml of CSF was withdrawn into sterile polypropylene tubes. The first 3 ml of the sample was sent to the local laboratory for analysis, which included analysis of cell count, protein, and glucose. CSF was then transferred in 0.2ml aliquots into pre-chilled polypropylene tubes, frozen immediately on dry ice, and stored at −80°C until analysis.

### CSF Measures

Aβ42, total tau, and phospho-tau (tau-p181) were measured using a Luminex-based INNO-BIA Alzbio3 assay, and Aβ40 was measured using the standard enzyme-linked immunosorbent assay (ELISA) according to manufacturer instructions, in each case using kits from Fujirebio (Malvern, PA; formerly Innogenetics, Belgium). We quantified CSF neurogranin using an in-house sandwich ELISA assay,^85^ sTREM2 using an in-house Meso Scale Discovery assay^86^. Baseline and follow-up samples were analyzed side-by-side on the same measurement plates, and intra-assay coefficients of variation were below 10%.

### Neuropsychological Evaluation

At baseline and after each diet, study participants completed assessments of immediate and delayed memory. Tests included the Free and Cued Selective Reminding Test (FCSRT),^87^ Story Recall (modification of the episodic memory measure from the Wechsler Memory Scale-Revised),^88^ and the Alzheimer’s Disease Assessment Scale-Cognitive (ADAS-Cog12).^89^ Cognition was assessed before and after each diet, and at follow-up. Different versions of selected tests were utilized to mitigate the impact of practice effects on cognitive performance.

### Statistical Analyses

Statistical analysis was conducted using R (4.1.3). Lipidomics data was log10 transformed prior to analysis. Linear mixed models (R package ‘lme4’) using baseline and follow-up samples from the MMKD and AHAD intervention arms were used to assess lipid changes with diet. P-values from the linear mixed model were derived using the package R package ‘lmerTest.’ An additional interaction term was added to assess differences in plasma lipid associations to dietary intervention based on clinical subgroups (cognitively normal or mild cognitive impairment, *APOE* □*4* status, or sex). P-values were adjusted for false discovery rate (FDR) using the approach by Benjamini and Hochberg^90^ separately for the lipid species (784) and classes (47).

To identify relationships between plasma lipids and CSF/cognition markers, the absolute differences (follow-up minus baseline) were calculated for each variable and lipid. Samples from the MMKD treatment arm were utilized in a Pearson correlation. Samples with missing CSF/cognition values were excluded from the specific analysis.

## Data Availability

Samples were provided by Wake Forest Alzheimer's Disease Research Center. Clinical data can be requested from the National Alzheimer's Coordinating Center (https://naccdata.org/).

## Acknowledgments and Funding

Samples were provided by Wake Forest Alzheimer’s Disease Research Center (WFADRC) supported by (P30AG049638) and associated laboratory staff, the Hartman Family Foundation, and the Roena B. Kulynych Center for Memory and Cognition Research. We would like to acknowledge the contributions of the Wake Forest Clinical and Translational Science Institute which is supported by the National Center for Advancing Translational Sciences (NCATS) through National Institutes of Health grant UL1TR001420. Additional support was provided by the Swedish Research Council and Swedish State Support for Clinical Research (ALFGBG) and laboratory technicians at Sahlgrenska University Hospital, Mölndal, Sweden.

Metabolomics data is provided by the Alzheimer’s Gut Microbiome Project (AGMP) or the Alzheimer’s Disease Metabolomics Consortium (ADMC) funded wholly or in part by the following grants and supplements thereto: NIA R01AG046171, RF1AG051550, RF1AG057452, R01AG059093, RF1AG058942, U01AG061359, U19AG063744 and FNIH: #DAOU16AMPA awarded to Dr. Kaddurah-Daouk at Duke University in partnership with a large number of academic institutions. A listing of AGMP Investigators can be found at https://alzheimergut.org/meet-the-team/. A complete listing of ADMC investigators can be found at: https://sites.duke.edu/adnimetab/team/. Dr. Huynh is supported by a National Health and Medical Research Council (NHMRC) investigator grant (1197190). JK and RB are supported by the National Institute of Aging of the National Institutes of Health under awards 1U19AG063744 and R01AG069901-01. RB is also supported by Alzheimer’s association award AARFD-22-974775. Dr. Zetterberg is a Wallenberg Scholar supported by grants from the Swedish Research Council (#2022-01018 and #2019-02397), the European Union’s Horizon Europe research and innovation programme under grant agreement No 101053962, Swedish State Support for Clinical Research (#ALFGBG-71320), the Alzheimer Drug Discovery Foundation (ADDF), USA (#201809-2016862), the AD Strategic Fund and the Alzheimer’s Association (#ADSF-21-831376-C, #ADSF-21-831381-C, and #ADSF-21-831377-C), the Bluefield Project, the Olav Thon Foundation, the Erling-Persson Family Foundation, Stiftelsen för Gamla Tjänarinnor, Hjärnfonden, Sweden (#FO2022-0270), the European Union’s Horizon 2020 research and innovation programme under the Marie Skłodowska-Curie grant agreement No 860197 (MIRIADE), the European Union Joint Programme – Neurodegenerative Disease Research (JPND2021-00694), the National Institute for Health and Care Research University College London Hospitals Biomedical Research Centre, and the UK Dementia Research Institute at UCL (UKDRI-1003).

## Conflict of interest

Dr. Kaddurah-Daouk in an inventor on a series of patents on use of metabolomics for the diagnosis and treatment of CNS diseases and holds equity in Metabolon Inc., Chymia LLC and PsyProtix. JK holds equity in Chymia LLC and IP in PsyProtix and is cofounder of iollo. JK holds equity in Chymia LLC and IP in PsyProtix and is cofounder of iollo. Dr. Zetterberg has served at scientific advisory boards and/or as a consultant for Abbvie, Acumen, Alector, Alzinova, ALZPath, Annexon, Apellis, Artery Therapeutics, AZTherapies, CogRx, Denali, Eisai, Nervgen, Novo Nordisk, Optoceutics, Passage Bio, Pinteon Therapeutics, Prothena, Red Abbey Labs, reMYND, Roche, Samumed, Siemens Healthineers, Triplet Therapeutics, and Wave, has given lectures in symposia sponsored by Cellectricon, Fujirebio, Alzecure, Biogen, and Roche, and is a co-founder of Brain Biomarker Solutions in Gothenburg AB (BBS), which is a part of the GU Ventures Incubator Program (outside submitted work).

## Notes

### Clinical Trial

NCT02984540

### Author Declarations

Samples were provided by Wake Forest Alzheimer's Disease Research Center. Clinical data can be requested from the National Alzheimer's Coordinating Center (https://naccdata.org/). The protocol was approved by the Wake Forest Institutional Review Board, and written informed consent was obtained from all participants and/or their study partners.

