## Supplementary Figures 1-3 for "Therapeutic potential of a modified Mediterranean ketogenic diet in reversing the peripheral lipid signature of Alzheimer’s disease"

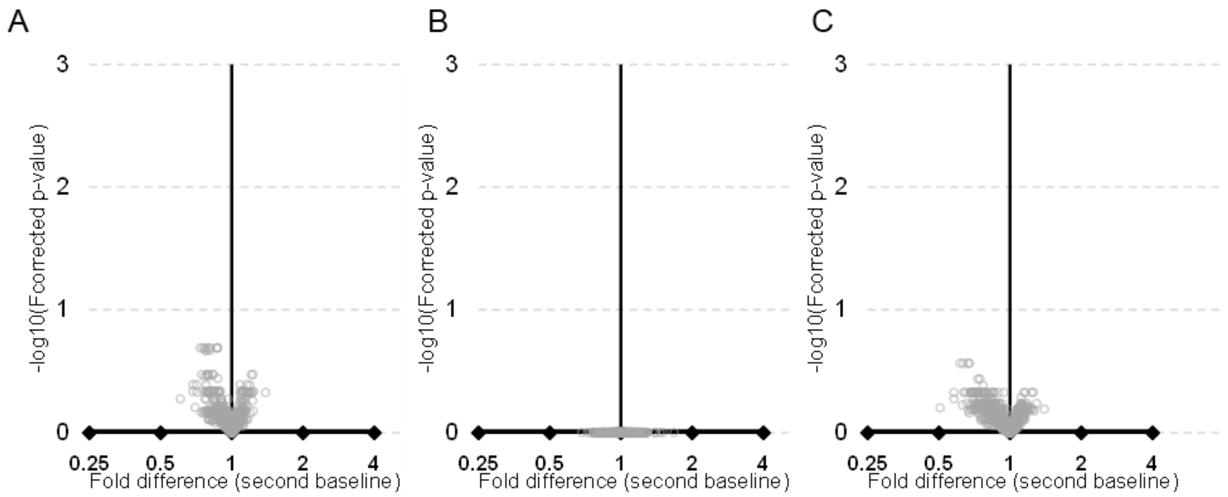


**Supplementary Figure 1 – Lipid differences post-washout.** Volcano plots generated from a linear mixed model to assess differences between baseline at the first and second diet intervention. **A -** Fold differences associated with any intervention **B -** Fold differences associated with starting on the MMKD intervention arm at baseline. **C -** Fold differences associated with starting in the AHAD intervention arm at baseline.

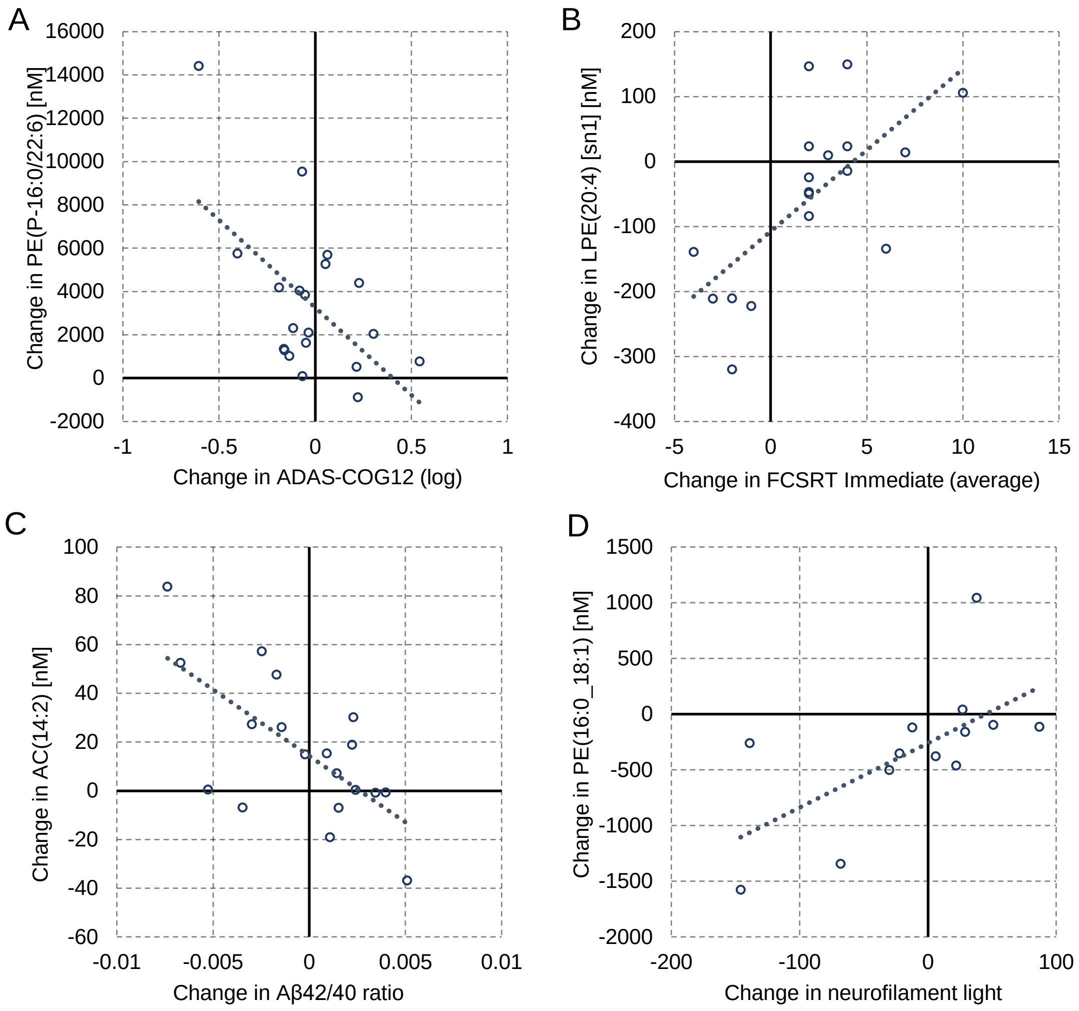


**Supplementary Figure 2 – Relationship between changes to the lipidome and clinical measures in response to the MMKD.** A – Inverse relationship between changes to ADAS-Cog12 and PE(P-16:0/22:6), B – Positive associations between FCSRT (immediate, average) and LPE(20:4), C – Inverse relationship between changes to AC(14:2) and changes to Aβ42/40 ratio, D – Positive relationship between PE(16:0_18:1) and neurofilament light.

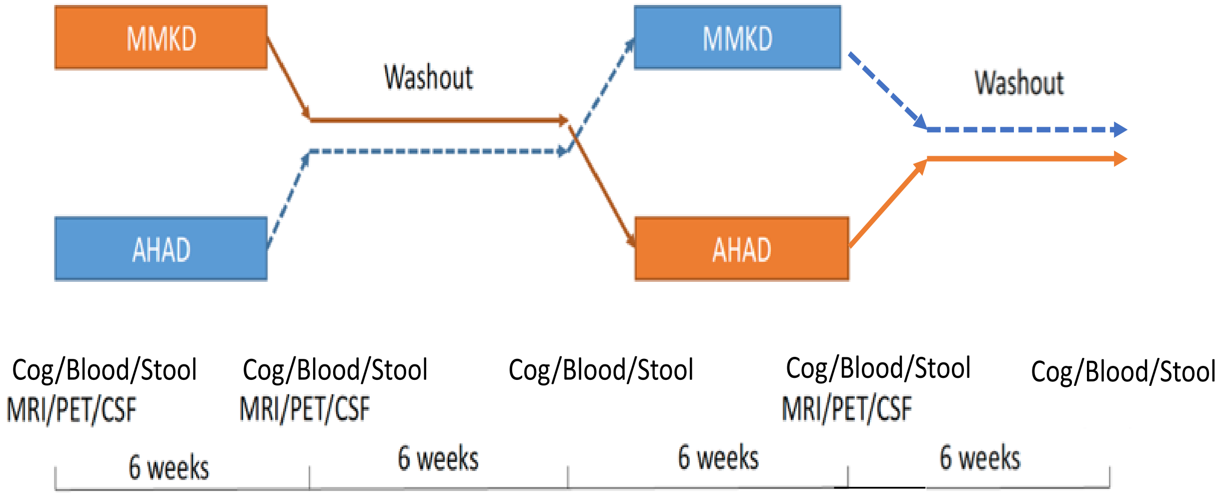


**Supplementary Figure 3 - Study Design.** The study used a randomized crossover design in which participants consumed either a MMKD or the control AHA diet for 6 weeks, followed by a 6-week washout period in which participants were instructed to resume their pre-study diet, after which the second diet was consumed for 6 weeks. Participants underwent LP/MRI before and after the first diet, and then following the second diet. Cognitive testing and blood/stool collection occurred before and after each diet.
